# Viral etiology of respiratory infections among patients at Adama Hospital Medical College, a facility-based surveillance site in Oromia, Ethiopia

**DOI:** 10.1101/2024.05.31.24308236

**Authors:** Bedado Dulo, Gamachu Hinsene, Ephrem Mannekulih

**Affiliations:** Adama Hospital Medical College, Department of Biomedical Sciences, Microbiology and Parasitology unity, Adama, Ethiopia; Adama Regional Laboratory capacity building, Microbiology, and Public Health expertise, Adama, Ethiopia; Adama Hospital Medical College, Department Public Health Adama, Ethiopia

**Keywords:** Severe Acute Respiratory Infection, Influenza, SARS-CoV-2, RSV, coinfection

## Abstract

**Background:** Acute viral origins account for around 80% of respiratory illnesses globally. The influenza virus, respiratory syncytial virus, coronavirus, adenovirus, and rhinovirus are the main viruses that cause these illnesses. All ages are susceptible to severe acute respiratory infections, which have a high rate of morbidity and mortality.

This study aims to determine the prevalence of viral etiology of respiratory infections among patients attending the Oromia Sentinel Surveillance Sites between July 2022 and April 2023.

**Methods:** A facility-based cross-sectional study design was employed. We followed the WHO case definitions for each patient with a severe acute respiratory infection. The throat-swab specimens were sent to the Adama Public Health Referral and Research Capacity Building Centre after being collected in viral transport media.

After that, the CDC Multiplex RT-PCR amplification procedures were applied to the specimens to detect the presence of viral RNA using CDC Real-Time reverse transcription PCR techniques. Data quality assurance was maintained. SPSS version 29 statistical software was used to compute all analyses. At 95% CI and P-value <0.05, inferential analysis was performed.

**Results:** The results of this study showed that out of three hundred twenty-two throat-swab specimens collected, 100% underwent testing. Eleven (28.2%) of the thirty-nine (12.9%) who tested positive for influenza were influenza B, twenty-five (89.3%) were influenza A (H3N2), three (10.7%) were influenza A (H1N1) pdm2009.

The rates of influenza positivity by age group were 58.9%, 25.6%, 5.1%, 5.1%, and 5.1% for children under five years old, 5–14 years old, 15–49 years old, 50–64 years old, and older than or equal to 65 years old.

Three hundred and twenty-two (100%), twenty-two (7.3%), and eleven (3.6%) of the specimens examined for severe acute respiratory infections proved positive for the RSV and SARS-CoV-2 viruses, respectively. Furthermore, of the severe acute respiratory infection specimens that tested positive for Respiratory syncytial virus, 91% were from under five age groups.

**Conclusion:** Children under five are at risk of co-infection with various viruses, potentially leading to epidemics and severe illnesses. A comprehensive approach to IPC measures is needed to reduce these risks.

## INTRODUCTION

Acute respiratory infection (ARI) is a significant global health concern, with influenza being the leading cause [1-2]. Influenza viruses are enveloped negative-stranded RNA viruses with segmented genomes containing eight gene segments [4]. They have a wide range of hosts, including humans, birds, pigs, horses, and marine mammals [3]. Seasonal epidemics are generally caused by types A and B, responsible for 3 to 5 million cases of severe acute respiratory infection (SARI), resulting in significant global morbidity and mortality every year and 290,000 to 500,000 deaths per year [5-6].

The severity of influenza viruses is mainly due to aggravations of underlying conditions, such as bacterial co-infection and secondary infection that synergize influenza viruses, leading to severe complications like respiratory distress syndrome (ARDS), respiratory failure, and death [7-9]. Seasonal influenza viruses evolve continuously and cause severe disease annually, particularly in the elderly, children, pregnant women, and people with underlying chronic conditions [10-12].

In Africa, influenza-related incidence data remain inadequate and outcome data are still very limited. Seasonality patterns of influenza in eastern Africa, including Ethiopia, have not been established [13-15]. Co-infection with various viral pathogens has been confirmed in a substantial number of patients with respiratory tract diseases, but the association between the occurrence of co-infection and substantially higher severity of disease is still unclear [16-21].

The influenza virus, causing significant illness and death, is a major cause of respiratory tract illnesses, hospitalization, and mortality, especially in infants and young children [22-23]. The COVID-19 pandemic has significantly impacted healthcare services worldwide [24-26]. Measures to reduce coronavirus transmission have also reduced the transmission of other respiratory viruses. However, the clinical outcomes of respiratory viral co-infections with coronavirus remain unknown. The Ethiopian Influenza sentinel surveillance system faced challenges, including weak follow-up of influenza-positive cases, high SARI patient enrollment, and limited data collection [27-28]. This study aims to identify influenza virus types and subtypes and determine their association with disease outcomes and socio-demographic variables.

## Materials and Methods

### Study area

The Adama Public Health Research and Referral Laboratory Center (APHRRLC), established in 1996, is located in Oromia Regional State, Adama City, and 100 km from Addis Ababa. Launched in 2003, it serves as a study site for Oromia severe acute respiratory infections Sentinel Surveillance reports.

### Study design and period

The studyconducted from July 2022 to April 2023, was a prospective hospital-based study utilizing data from severe acute respiratory infection sentinel surveillance.

### Population

#### Source population

The patient list for the 2022/2023 season, from July 2022 to April 2023, includes all those attending Adama Hospital Medical College.

#### Study population

Sentinel surveillance staff collect data on patients with severe acute respiratory infection, following the World Health Organization case definition, as part of epidemiological surveillance.

#### Sample size determination and sampling technique

During the study period, 322 sample sizes were included from sentinel surveillance sites, following the World Health Organization case definition for severe acute respiratory infection.

#### Inclusion criteria

Legible cases meeting the World Health Organization case definitions for severe acute respiratory infection from the catchment of Adama Hospital Medical College sentinel surveillance were included.

#### Exclusion criteria

The patients were included in the Severe acute respiratory infection case definitions protocol guideline. Those patients did not meet the protocol, and or refused to provide the sample excluded.

### Study variables

#### Dependent variable

The positivity rate of influenza and co-infection with severe acute respiratory syndrome -CoV -2 or Respiratory syncytia viruses were the dependent variables

#### Independent variables

The patient’s demographic data, including age, sex, address, occupation, pre-existing conditions, vaccinations, influenza exposure, travel history, and other factors, were considered independent variables..

### Operational definition

#### Who’s severe acute respiratory infection case definitions

A severe acute respiratory infection case is a patient over 5 years old with sudden fever, cough, sore throat, shortness of breath, hospitalization, or a child under 5 years [37].

**Co-infection:** Infection with Influenza and either coronavirus or Respiratory syncytial virus.

**Co-circulation:** The availability of various viruses together in the community either infecting a patient separately or in the form of co-infection.

#### Data collection tools and technique

A severe acute respiratory infection case-based data collection form was used to gather demographic and clinical data from adult patients and parents. Symptoms, comorbidities, and samples were collected from nasopharyngeal swabs after clinical examination. The samples were transported to the Ethiopian Health Institute and APHRRLC for diagnosis, and stored at -20oc until testing. The data was then sent to other laboratories for further testing.

#### Data quality assurance

Data quality assurance was maintained through orienting data collectors and supervisors on data collection, sample collection, and transportation. Supervisors check severe acute respiratory infection case-based data forms for completeness, accuracy, and timeliness before sending them weekly to APHRRLC or EPHI. Data arrives at laboratories, is checked for completeness, accuracy, and timeliness, and is cleaned before being entered into Epi info version 7.2.5 and exported to SPSS version 26 for analysis.

#### Data management and analysis

The study used descriptive statistics, Pearson chi-square, and Fisher’s exact tests to compare categorical variables and two-sample t-tests for continuous variables. Seasonality and trends of influenza type and subtype were evaluated using an epidemiologic curve. The positivity rate for influenza was calculated, and differences in positivity were tested based on infection severity and age group.

#### Ethical review

In Ethiopia, Oromia region is one of the National Influenza Sentinel surveillance sites for Severe Acute Respiratory infections studies framed with protocol and supervised by the Ethiopian Public Health Institute.

The Adama Public Health Research and Referral Laboratory Center conduct a routine surveillance patients fulfilling the nation protocol to address the national target. Ethical clearance was obtained from the Oromia Health Bureau Research Ethical Review Board with letter reference no/Lkk: BFI/HBTH/ 1-16/4035 written on date 21/3/2015 E.C. Unwritten consent was obtained from parents for underaged patients.

#### Dissemination of study the findings

The study aids stakeholders in decision-making on interventional prevention against seasonal influenza, including vaccination, reducing severe acute respiratory infection incidence, hospitalization, and case fatality, and will be presented at health institutes and conferences.

## Results

### Demographic and clinical Characteristics of study subjects

A study enrolled 302 patients with severe acute respiratory infections, with a mean age of 9.6 years and a median age of 3.1 years. Males accounted for 56.6% of the patients, with comorbidities including liver disease, chronic heart disease, and heart disease. Symptoms included difficulty breathing, sore throat, and runny nose. Only 3 patients were vaccinated [Table 1].

**Table 1:** Demographic and clinical Characteristics of study subjects at Adama Hospital Medical *College’s* Influenza Sentinel Surveillance Site from July 2022 through April 2023.

| Characteristics |  | Frequency | Percent | 95% Confidence Interval |  |
| --- | --- | --- | --- | --- | --- |
|  |  |  |  | Lower | Upper |
| Age Category | 0-1.9 | 89 | 29.5 | 24.5 | 35.1 |
|  | 2-4 | 91 | 30.1 | 24.8 | 35.1 |
|  | 5-14 | 74 | 24.5 | 19.9 | 29.5 |
|  | 15-49 | 32 | 10.6 | 7.3 | 14.2 |
|  | 50-64 | 9 | 3.0 | 1.3 | 5.0 |
|  | >=65 | 7 | 2.3 | 1.0 | 4.3 |
|  | Total | 302 | 100.0 | 100.0 | 100.0 |
| Sex | Male | 171 | 56.6 | 51.0 | 62.3 |
|  | Female | 131 | 43.4 | 37.7 | 49.0 |
|  | Total | 302 | 100.0 | 100.0 | 100.0 |
| Zone | Adama Town | 46 | 15.2 | 11.3 | 19.5 |
|  | Arsi | 75 | 24.8 | 20.2 | 30.1 |
|  | Borana | 1 | .3 | .0 | 1.3 |
|  | East Shoa | 179 | 59.3 | 53.6 | 64.9 |
|  | Harerge | 1 | 0.3 | - | 1.0 |
|  | Total | 302 | 100.0 | 100.0 | 100.0 |
| Comorbidities | No known condition | 290 | 96.0 | 93.7 | 98.3 |
|  | LD | 2 | 0.7 | - | 1.7 |
|  | CLD | 8 | 2.6 | 1.0 | 4.6 |
|  | CHD | 1 | 0.3 | - | 1.0 |
|  | HD | 1 | .03 | .- | 1.0 |
|  | Total | 302 | 100.0 | 100.0 | 100.0 |
| Difficulty of Breathing | No | 14 | 4.6 | 2.3 | 7.3 |
|  | Yes | 288 | 95.4 | 92.7 | 97.7 |
|  | Total | 302 | 100.0 | 100.0 | 100.0 |
| Sore Throat | No | 20 | 6.6 | 4.0 | 9.6 |
|  | Yes | 282 | 93.4 | 90.4 | 96.0 |
|  | Total | 302 | 100.0 | 100.0 | 100.0 |
| Runny Nose | No | 34 | 11.3 | 7.9 | 14.9 |
|  | Yes | 268 | 88.7 | 85.1 | 92.1 |
|  | Total | 302 | 100.0 | 100.0 | 100.0 |

### Prevalence of Influenza Virus, Types, and Subtypes

A total of 302 samples were tested for influenza using multiplex RT-PCR, with 39 positive (12.9%) of the samples being positive for different types and subtypes. Influenza type A accounted for 28 (71.8%) cases, with A (H1N1) pdm2009 and H3N2 contributing 89.3% and 28.2% respectively. Males accounted for 69.2% of positive cases, while A (H3N2) contributed 89.3%, with 36%, 28%, and 20% attributed to age categories. Influenza type B distribution was 36.4%, 27.3%, and 27.3%, with males contributing 72.7% [Table 2].

**Table 2:**
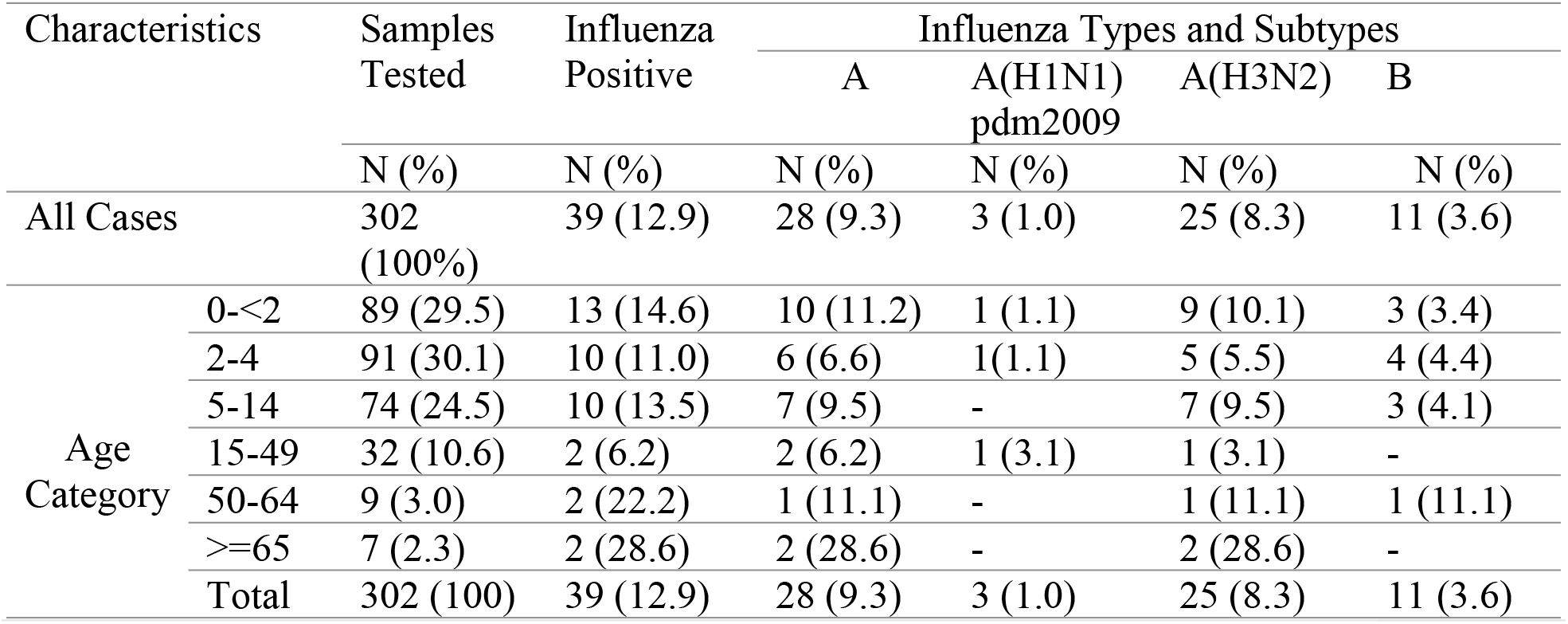
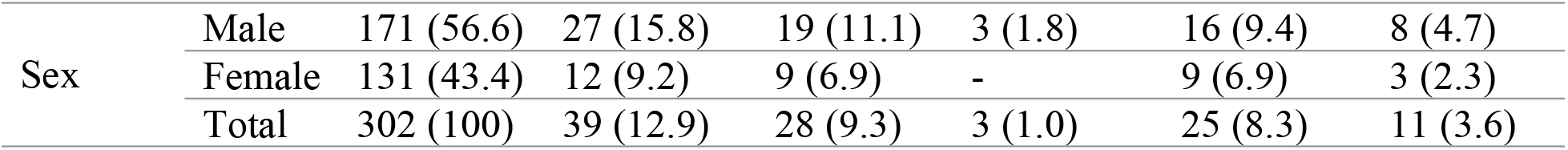
Prevalence of Influenza virus, Types, and Subtypes among Study Subjects at Adama Hospital Medical *College’s* Influenza Sentinel Surveillance Site from July 2022 through April 2023.

### Prevalence of severe acute respiratory syndrome, and respiratory syncytial viruses

Out of 302 Severe acute respiratory infection specimens, 22 (7.3%) and 11 (3.6%) were positive for Severe acute respiratory syndrome, and Respiratory syncytial viruses. Age categories contributed to severe acute respiratory syndrome positivity, with males contributing 63.6%. Respiratory syncytial prevalence was also influenced by age, with males contributing 63.6%. Both positive cases were found in under 2 years and 2-4 years old [Table 3].

**Table 3:** Prevalence of Severe acute respiratory syndrome, and Respiratory syncytial viruses among study subjects at Adama Hospital Medical College’s Influenza Sentinel Surveillance Site from July 2022 through April 2023.

| Characteristics |  | Samples Tested | Severe acute respiratory syndrome Positive | Respiratory syncytial virus Positive |
| --- | --- | --- | --- | --- |
|  |  | N (%) | N (%) | N (%) |
| All Cases |  | 302 (100%) | 22 (7.3) | 11 (3.6) |
| Age Category | 0-<2 | 89 (29.5) | 10 (11.2) | 5 (5.6) |
|  | 2-4 | 91 (30.1) | 6 (6.6) | 5 (5.5) |
|  | 5-14 | 74 (24.5) | - | 1 (1.4) |
|  | 15-49 | 32 (10.6) | 3 (9.4) | - |
|  | 50-64 | 9 (3.0) | 1 (11.1) | - |
|  | >=65 | 7 (2.3) | 2 (28.8) | - |
|  | Total | 302 (100) | 22 (7.3) | 11 (3.6) |
| Sex | Male | 171 (56.6) | 14 (8.2) | 7 (4.1) |
|  | Female | 131 (43.4) | 8 (6.1) | 4 (3.1) |
|  | Total | 302 (100) | 22 (7.3) | 11 (3.6) |

## Discussion

The study found that the proportion of influenza-positive specimens among enrolled severe acute respiratory infection patients at Adama Hospital Medical College was 13.3% from July 2022 to April 2023. The peak in influenza positives was observed during the winter months, with a distinct peak in December 2022. The proportion of influenza A and B was higher than previous reports in Ethiopia 3%[40], and Egypt, but lower than previous reports in Egypt, Ethiopia, and Pakistan[42, 43, 47, 48]. However, our current study result (13%) is similar to reports from Beijing(China), Indonesia, Malawi, Kenya, Central China, and Zambia 16.6%, 12.1%, 14.5%, 10%, 13%, and 16.3%[7, 33, 34, 36, 41, 45] respectively. Influenza A (H3N2) was attributed to 25 (89.3%) cases, while A (H1N1) pdm2009 was lower than previous reports. Influenza coinfection with Severe acute respiratory syndrome -CoV-2 was 12.8%, with higher coinfection in influenza subtype A. Co-circulation and co-infection of A(H3N2), A(H1N1) pdm2009, Influenza B, and Severe acute respiratory syndrome-CoV-2 viruses were common in under 5 years old children. During the study, 8 cases of co-infections were detected, with 5 cases of severe acute respiratory syndrome, and influenza and 3 cases of respiratory syncytial virus and influenza. Influenza coinfection with Respiratory syncytial virus was not discussed due to the small number of cases. Other studies reported variable prevalence and clinical outcomes of influenza coinfection with severe acute respiratory syndrome -CoV-2, with differences in regions and geographical locations.

## Conclusion and Recommendation

Children under five age included in this study, are at risk of co-infection with various viruses, potentially leading to epidemics and severe illnesses. A comprehensive approach to Infection Prevention Control (IPC) measures is needed to manage these risks. Multiplex RT-PCR testing for influenza and severe acute respiratory syndrome -CoV-2 is necessary, and public health strategies like vaccination and mask use during high respiratory virus circulation are recommended. Further studies with larger sample sizes are also warranted to study the associated risk factors of coinfection.

## Data Availability

All data produced in the present study are available upon reasonable request to the authors.

## Acknowledgments

The authors express gratitude to the Oromia Health Bureau, Adama Public Health Research & Referral Laboratory Center, Adama Hospital Medical College, Mojo Hospitals, and study participants for their support and volunteerism.

